# Variable selection for competing risk regression models: recommendations for analyzing data from epidemiological studies

**DOI:** 10.1101/2024.11.25.24317882

**Authors:** J. Mullaert, Sandra Schmeller, Peter C. Austin, A. Latouche

## Abstract

When fitting competing risks regression models, a variety of variable selection methods exist, including backward selection on the subdistribution hazard, on the cause-specific hazards, and penalized methods. However, a benchmark study comparing these different procedures is lacking.

We conducted an extensive simulation study to compare three variable selection procedures in terms of both model selection ability and predictive accuracy. 5120 datasets were simulated in various conditions aiming at being representative of real applications in clinical epidemiology. Results show that the performance of backward selection procedure can be affected by implementation choices. Even for scenarios with a high numbers of events per variable (EPV), the true model is rarely identified by any of the selection procedures. Survival predictions were assessed with time-dependent AUC and show similar performances for all methods. We also provided an application on stem cell transplanted patients in hematology.

We concluded that the identification of the true model in competing risk regression is a very difficult task, and suggest some recommendations to analysts: (1) to report event per variable for the event type of interest and (2) to use multiple methods to deal with model uncertainty and avoid implementation pitfalls.

## 1. Background

In clinical epidemiology, performing variable selection for a regression model (e.g., linear, logistic or Cox model) is a very common task. The main purpose is to identify clinical variables that are associated with the outcome and quantify their strength of association after adjusting for other confounding factors. The resulting multivariable model can also be used for making predictions. Many authors described good and bad practices for this task (1–3) and the development of new methods and algorithms for performing variable selection with diverse outcome types (e.g. count data, constrained quantitative data, time-to-event data) is still an active field of research (4–6). While penalized or resampling methods are increasingly popular for performing variable selection, stepwise procedures are still often used (despite being highly criticized), as well as building a multivariable model based on variables significantly associated with the outcome in the univariable analysis, or building a multivariable model based only on clinical considerations.

Competing risks occur when multiple events can arise and the occurrence of one precludes the occurrence of others. When modeling competing events, methods for variable selection have been less studied, for at least two reasons. First, two different families of competing risk models exist: the cause-specific hazard (CSH) is the natural extension of the hazard function, whereas the subdistribution hazard (SDH) preserves the one-to-one relation with the cumulative incidence function (7). The proportional hazard assumption can be made on the CSH or the SDH, but not both (8,9), although it is possible to have proportionality on one CSH and the corresponding SDH (10). In addition, with a continuous predictor variable, the hypothesis of proportionality on SDH cannot hold simultaneously for both event types (11). This co-existence of two possible parametrizations, each with their own interpretation and methodological pitfalls presumably contributed to the problem of variable selection in this context being less studied. Another reason comes from the additional complexity of this problem because two risks are simultaneously modelled. In the CSH parametrization, a variable can be included either as a predictor variable in the model for the first event type, for the second or for both. Any model misspecification, even on the second event type only, leads to a different cumulative incidence function (CIF) for the first event type because both CSH contribute to the definition of the all-cause hazard. Therefore, a higher number of candidate models is to be considered with respect to standard Cox regression, even if only the first competing risk is of clinical interest. This remark also applies when the objective of these models is to make predictions: both CSH models have to be correctly specified. This problem may be seen as less salient for the SDH parametrization because only the event of interest is usually modelled as it is enough to derive the CIF in this context. However, although this strategy is correct in theory, it strongly relies on the hypothesis of proportionality of SDH which may not be true in practice and may result in model misspecification and biased estimates (12).

These particularities, specific to the competing risk setting, possibly explain the paucity of recommendations on how to build a multivariable model for competing risk regression models (13), despite being available for other regression models (14). As a result, clinical papers investigating the association between potential risk factors and a clinical outcome mostly use heterogeneous and possibly suboptimal methods. Among them, backward elimination has been heavily criticized, despite being still widely used in clinical epidemiology (15,16). The main problem with backward elimination is that test statistics of the final model are biased and therefore cannot, in general, be used for inference. However, when the number of observed events per variable is high (>100), the method could arguably be used for variable selection and making predictions (3). This is why some authors proposed such algorithm in the presence of competing risks and apply the backward elimination procedure on either both CSH (17) or the SDH of the event of interest (18). As an alternative to stepwise methods, penalized regression performs variable selection by adding a penalty term based on the L1 norm to the partial likelihood, which forces some coefficients to be exactly 0. The penalization level has to be tuned by cross-validation in order to maximize some efficiency criterion (e.g., AUC).

Our objective was to compare three existing and widely used algorithmic procedures for variable selection in the presence of competing risks. Our main goal was to assess their ability to (i) identify the true underlying model and (ii) make accurate survival predictions, as quantified by the time dependent area under the curve (AUC). The scope of this paper is restricted to clinical epidemiology, where a reasonable number of events (typically 100-1000) is involved with a limited number of variables (typically 10-20). It does not cover the case of high-dimension data (e.g., from omics studies) or analysis of big databases, although it can apply to such data after a dimension reduction procedure being performed.

The rest of this paper is organized as follows. Section 2 is devoted to the presentation of the design of the simulations and statistical analyses. In section 3, results of the simulation study are reported. In Section 4, a real data application for the prognosis after hematopoietic stem cell transplantation is described. Finally, in Section 5 we provide some discussion elements.

## 2. Methods

### Simulation study

For the sake of simplicity, a simulation study with two competing events was considered. Datasets of different sample sizes (100, 200, 400 and 800) were generated, with a model assuming proportional cause-specific hazards or proportional subdistribution hazard for event type 1 (models 1 and 2 respectively).

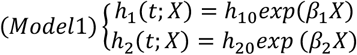

According to model 1, the two cause-specific hazards denoted *h*_1_ and *h*_2_ are specified with constant baseline hazards *h*_10_ and *h*_20_ and covariates have a proportional effect on each cause-specific hazards. The procedure to generate event times and types from this model is described in (19). Model 2 is specified with cumulative incidence functions (CIF) *F*_1_ and *F*_2_, such that the effect of covariates is proportional on the subdistribution hazard of risk 1.

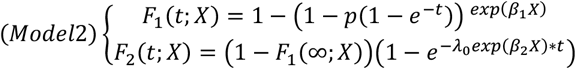

Of note, it is not possible to ensure a proportional effect on both subdistribution hazards. Therefore, the shape of *F*_2_ is calibrated such that the sum of both CIF tends to 1 as time goes to infinity, through the expression 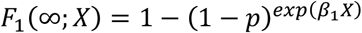,where *p* denotes the limit *F*_1_(+∞;0). For this model, as both CIF are fully specified, the procedure to generate data is more straightforward than for model 1. Let z be a realization of a uniform distribution over (0,1). If *z* < *F*_1_(+∞; *X*), then the event type is 1 and the event time is 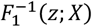. If *z* > *F*_1_(+∞; *X*), then the event type is 2 and the event time is 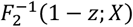.

In both models, *X* denotes a vector of 12 or 24 independent covariates. This design is inspired from the simulation section of (20): the first six components follow a standard normal distribution, and the next six components follow a Bernoulli distribution with parameter *p* = 0.5. In scenarios with 12 covariates, the vector of coefficients *β*_1_ and *β*_2_ are as follows:

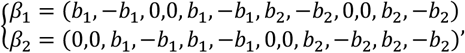

where *b*_1_ and *b*_2_ are effect sizes. Under the null hypothesis of no association between covariate and event times, we have *b*_1_ = *b*_2_ = 0. Another simulation is performed under the alternative hypothesis *b*_1_ = 0.9 and *b*_2_ = 0.4. Therefore, under the alternative hypothesis, covariates 1, 5, 7 and 11 (resp. 2, 6, 8 and 12) are positively (resp. negatively) associated with event 1, whereas covariates 3, 5, 9 and 11 (resp. 4, 6, 10 and 12) are positively (resp. negatively) associated with event 2. In some simulation scenarios, 12 additional covariates following the standard normal distribution, with no association with event times were simulated. Finally, four scenarios (S1 to S4) involving different baseline hazards and censoring rate were considered. Details about the parameters used in the simulation are given in supplementary table **S1** and **S2**. These parameters were chosen to be representative of real applications: a ratio between baseline hazards or cumulative incidences equal to 1 or 4 with small or moderate censoring in model 1 and 2 respectively.

To sum up the simulation settings, the following factors were allowed to vary: (i) 4 different sample size (100, 200, 400 and 800), (ii) 2 theoretical models for data generation (model 1, proportional CSH or 2, proportional SDH), (iii) 2 possible association scheme (null hypothesis *b*_1_ = *b*_2_ = 0 or alternative hypothesis *b*_1_ = 0.9 and *b*_2_ = 0.4), (iv) 2 different dimensionality for the vector of covariates (size 12 or 24), (v) 4 scenarios of baseline hazards and censoring rate (see table **S1**). We thus considered 128 different scenarios. For each scenario, we simulated 200 datasets for model training and 200 of the same size for external validation, resulting in 51200 datasets being simulated.

### Variable selection procedures

For each training dataset, three variable selection procedures using R packages were performed. The first procedure corresponds to a CSH model and the other two to a SDH model. These three procedures were applied to each simulated dataset (irrespectively of the model used for simulation) to evaluate the robustness for model misspecification. This choice also has the advantage of not favoring any parametrization because the true data generating mechanism is, in real life, not known.

First the *SelectCox* function from the package *pec* performs backward selection on a Cox model to minimize the Akaike Information Criterion (AIC). In this study, it was used sequentially for fitting two cause-specific hazard models according to the following procedure: *β*_1_ was estimated by considering patients experiencing event 2 as censored observation for event 1. This leads to a model with covariates associated with the cause-specific hazard of event 1. The same procedure was performed for the second event type.

The *SelectFGR* function from the package *crrstep* (18) performs backward selection for a Fine-Gray subdistribution hazard model, i.e., a semi parametric proportional subdistribution hazard model. This function provides an estimation of *β*_1_ only. Importantly, the practical implementation of the backward selection is not the same as the *SelectCox* method in that it relies on the *stepAIC* function of *MASS* package, whereas *SelectCox* uses the *fastbw* function from package *rms*. The fast backward algorithm only fits the full model and then derives approximation of Wald statistics of submodels without fitting them. As a consequence, the computational burden is low, but this approximation is less valid if the number of deleted variables is large. Of note, both functions *SelectFGR* and *SelectCox* use AIC as stopping rule.

The *crrp* function from the package of the same name (21) performs a penalized estimation of the proportional subdistribution hazard model with a LASSO penalty, minimizing the Bayesian Information Criterion (BIC). This method provides an estimate of *β*_1_ only. Of note, the *crrp* package has recently been removed from the CRAN repository due to its maintainer not being reachable, but remains available for download as an archive.

### Performance metrics

Under the null hypothesis of no association (i.e., all the regression coefficients being equal to zero), the model resulting from the variable selection procedure should be empty. Therefore, the number of wrongly selected variables was pooled across simulation replicates and expressed as a percentage of the total number of possible covariate (i.e., 12 or 24 depending on the scenario). The ratio corresponds to the probability of one covariate being wrongly present in the final model and is often reported as a false discovery rate (FDR).

Under the alternative hypothesis, the reported metric is the number of variables correctly selected (i.e., the number of selected variables among variables truly associated with the event risk), expressed as a percentage of all truly associated variables (i.e., 8 per simulation replicate). For example, if the selection procedure yields 3 variables truly associated to be selected (out of 6) and 2 variables not associated among 6, but selected anyway, the reported metric is 3/6=50%. We also considered the selection probability of variables that are not associated with the outcome to report by considering only the 12 last components of the covariate vector in scenarios with 24 covariates. For example, if the selection procedure yields 3 variables to be selected (out of 12), the reported metric is 3/12=25%. Of note, this metric does not consider the 12 first components of the covariate vector because these variables are all associated with one risk (either the first or the second) and this could lead to misleading results in the case of model misspecification, e.g. when the simulation is on CSH and the estimation on SDH. For the sake of clarity, all graphs under the null or alternative hypothesis are provided as nested loop plots (22).

As the resulting model can be used to obtain predictions which are of interest in certain applications, the predictive performances of the estimated models were assessed with the time dependent area under the curve (AUC), a measure of model discrimination, and scaled Brier score (sBS) for model calibration. Predictions are obtained either using the SDH of the event of interest, or both CSH if available. The time-dependent cumulative/dynamic AUC between time 0 and the median time of events occurrence were obtained from the *Score* function of the *RiskRegression* R package (23), on external validation samples. Similarly, the scaled Brier score corresponds to the ratio between (1) the Brier score of the considered model at the same median time as above and (2) the Brier score of a null model based on nonparametric survival estimate without any covariate. Since the Brier score corresponds to a quadratic error, high Brier score and high scaled Brier score mean that the model calibration is worse. Empirical mean of both AUC and sBS, 2.5% and 97.5% empirical quantiles were calculated and reported for each estimation methods, as well as for the true model that generated the data.

As a sensitivity analysis, we also investigated how our results on selection probability, discrimination and calibration can be altered by the use of BIC as a stopping rule in the backward selection procedure on subdistribution hazards.

## 3. Results

### False discovery rates under the hypothesis of no association

Results from the simulation under the null hypothesis of no association are reported in **Figure 1**. Backward selection based on the CIF of event 1 under proportional SDH assumption yielded a high number of variables being wrongly selected (around 15-20% across all scenarios), whereas backward selection on both CSH yields almost no false discoveries. The highest values of FDR occur in scenarios with small numbers of events per variable (EPV). This high FDR cannot be attributed to model misspecification since it is also observed when data were simulated under proportional SDH (i.e., model 2). Indeed, a modification of the SelectCox function enforcing the use of naïve backward elimination instead of fast backward algorithm yields a similar FDR as SelectFGR. Conversely, using fast backward elimination in SelectFGR results in almost no variable being selected (data not shown). The difference of FDR between the two methods is therefore only due to different practical implementations of the backward elimination algorithm.

**Figure 1:**
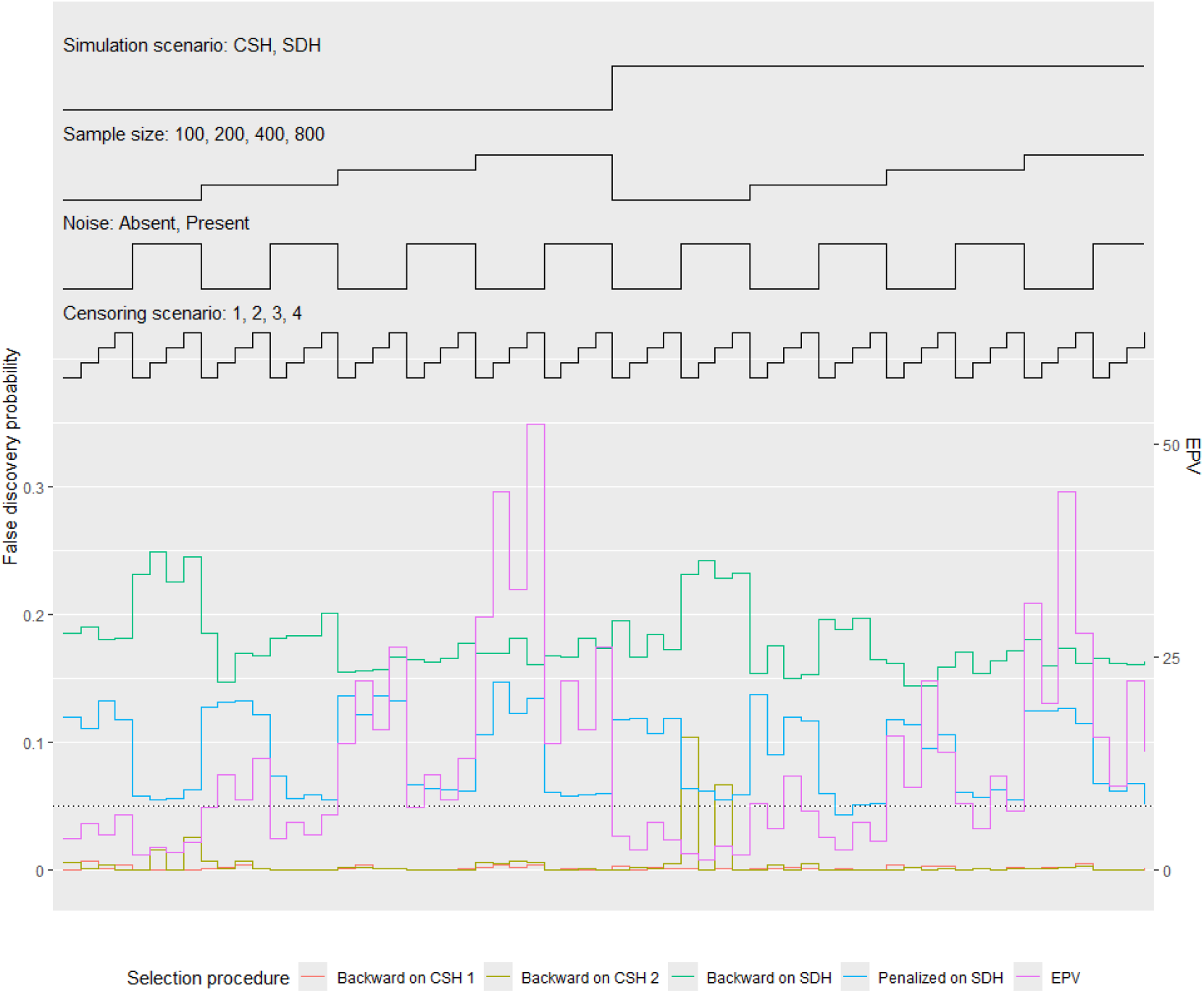
False discovery rate (left axis) and EPV (right axis) for selection procedures under the null hypothesis according to sample size, presence/absence of noise, censoring scenario and simulation scenario (64 scenarios).

Penalized selection on the SDH yields FDR ranging from 0.05 to 0.15, depending on the scenario considered. Interestingly, the highest FDR were obtained for scenarios with high EPV, suggesting that penalized selection is more reliable in a high-dimensional setting. In addition, high FDR is observed for the backward selection on CSH for event 2, in situations where the sample size is small and the censoring scenario yields a very small number of patients experiencing this event.

### Selection probability in the presence of non-null associations

Figure 2. reports the probability for an associated variable to be present in the final model for all tested selection methods. As expected from the results under the null hypothesis of no association, the probability for a truly associated variable to be selected for the final model is higher for the backward selection on CIF, than for other methods. As expected, this probability also increases with sample size and EPV, and is higher when the model is correctly specified (i.e. simulated with proportional SDH for selection methods on SDH and conversely for CSH).

**Figure 2:**
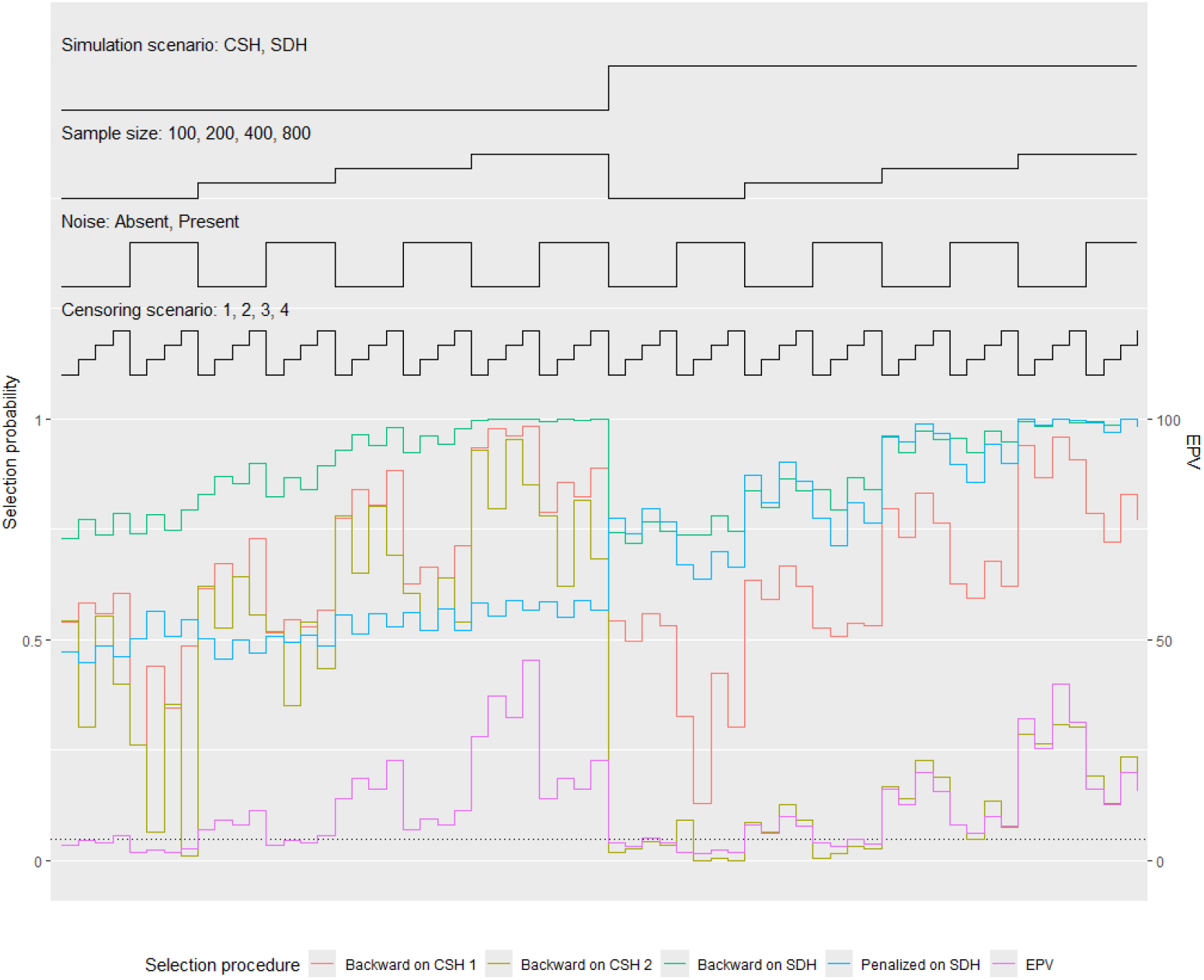
Selection probability (left axis) and EPV (right axis) for selection procedures under the alternative hypothesis according to sample size, presence/absence of noise, censoring scenario and simulation scenario (64 scenarios).

When data were generated under the proportional SDH model (model 2), 8 variables have to be simultaneously selected in order to get the true underlying model. At the same time, the 4 remaining variables must not be selected. This is why, except for scenarios with a high sample size and the penalized selection, the true model will almost never be identified. The situation is even worse in the case of data generated under the proportional CSH model because the identification of the true underlying model requires the selection of 16 associated variables for both risks and the absence of 8 unassociated variables. In this context, and considering observed selection probabilities, the selection procedure will almost never identify the true model.

**Supplementary Figure S3** reports the selection probability of covariates not associated with the outcome, while some other variables show associations with the outcome. Consistently with findings under the null hypothesis, the specificity is high for the backward selection procedure on CSH with almost all the selection probabilities below 5%. On the contrary, selection probabilities are higher for both selection methods on SDH, irrespectively of the model used for data generation.

### Prediction performances

The prediction performances and its variability for the different selection procedures and for the true model, when data are generated under model 2 (proportional SDH) with 24 covariates, scenario **S4** and N=200. **Figure 3** (left) shows the mean discrimination, evaluated on an independent validation dataset, are slightly lower than the one achieved by the true model and the variability is slightly larger. It is striking how small the differences are, with respect to the results of **Figures 1-2**. In other words, although the correct data generating model cannot be identified, the predictive performances, as measured with AUC, can be satisfactory. The same remark also applies to **Figure 3** (right), which presents the results of calibration.

**Figure 3:**
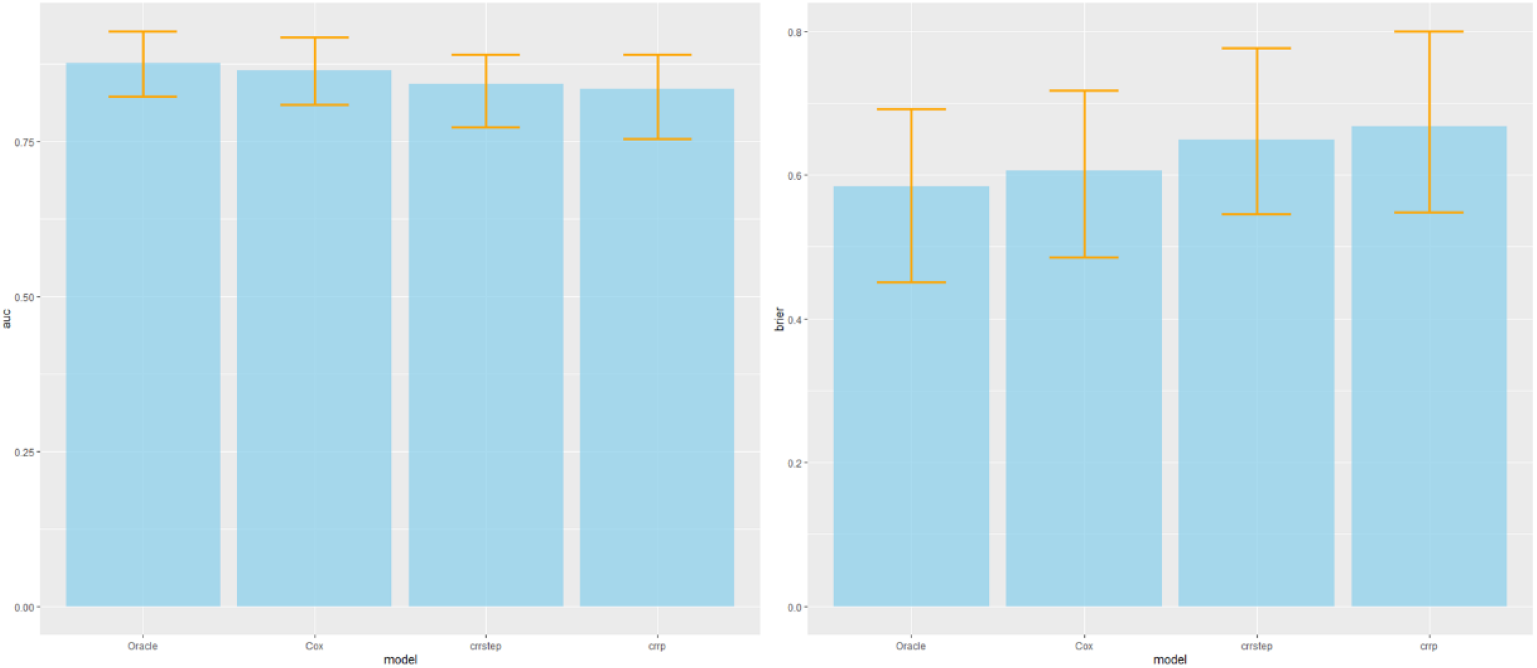
Discrimination (AUC) and calibration (scaled Brier score) performances of the studied selection methods on 100 simulated datasets with a proportional SDH model, N=200, 24 covariates and censoring scenario S4. Oracle designates the true model. Yellow lines indicate 95% confidence intervals.

The non-default use of BIC as a stopping rule in the backward selection procedure on the subdistribution hazard has been investigated on the same scenario as a sensitivity analysis. As expected, the use of BIC is associated with a lower selection rate for truly associated variables (70% and 56% for AIC and BIC respectively) and also for non-associated variables (33% and 15% for AIC and BIC respectively). The use of BIC has almost no impact on AUC (0.83 and 0.82 for AIC and BIC respectively) and scaled Brier score (71.2% and 71.3% for AIC and BIC respectively).

## 4. Application to real data

An allogeneic hematopoietic stem cell transplantation is often the only curative treatment for acute myeloid leukemia (AML). The success of a transplantation is often measured with disease free survival, but it is also important to distinguish the type of event: a relapse or death w/o prior relapse. Observing a relapse means that the transplantation failed whereas death w/o prior relapse can occur due to the transplantation as well as other comorbidities or an infection. Several risk factors can influence the outcome of a transplantation. On the one hand it is important to know which covariates influence the outcome and on the other hand the aim is to find the model with the best predictive performance. In this section we compare the three selection algorithms in the application to data from the German registry for stem cell transplantation (24), which has been conducted in accordance with the Declaration of Helsinki, and which received approval 226/17 and 141/19 from the ethical board of the Ulm University (Clinical trial number: not applicable). Informed consent had been retrieved prior to reporting.

Data from 3629 adult patients who were transplanted with an AML in Germany between 2016 and 2019 were available. The median follow-up time until an event or censoring was 9 months (0-65 months). The patient and covariable characteristics are shown in **Table 1**.

**Table 1:** Clinical characteristics of patients with hematopoietic stem cell transplantation (N=3629)

| Variable | Analyzed sample (N=3629)<br>mean (IQR) or N (%) |
| --- | --- |
| Patient Age (y) | 55 (49-65) |
| Donor Age (y) | 37 (26-46) |
| Type of donor: |  |
| - Identical sibling | 737 (20) |
| - Others | 2892 (80) |
| Sex combination: (missing=4) |  |
| - Male patient, female donor | 479 (13) |
| - Other | 3146 (87) |
| CMV mismatch (missing=62) | 1039 (29) |
| No previous autologous transplant | 3604 (99) |
| Karnofsky Performance Status<br>(missing=161) |  |
| - Good | 3196 (92) |
| - Poor | 272 (8) |
| Disease stage: |  |
| - Early | 1999 (55) |
| - Intermediate | 361 (10) |
| - Advanced | 1269 (35) |
| Myeloablative conditioning<br>(missing=126) | 1883 (52) |
| Total body irradiation (missing=24) | 690 (19) |
| Antithymocyte globulin (ATG) | 2517 (69) |
| Outcome |  |
| - Relapse | 973 (37) |
| - Death | 611 (17) |
| - Censored | 2045 (56) |

**Table 2** shows the regression coefficients for each endpoint and each selection procedure. On the cause-specific level, more covariables with a significant effect are selected for the event of death. The selection procedures for the subdistribution hazards selected the same covariables (patient age and disease stage) for endpoint relapse. The regression estimates only differ because of the penalty term implemented in *crrp*. For endpoint death the *crrp* -selection procedure chooses one more covariable compared to the *selectFGR*-procedure.

**Table 2:** CS_HR_ for the Cox model and the SD_HR_ subdistribution model after fitting the three variable selection procedures. A star (*) marks a significant difference to 1 with a significance level of 5%.

| Variable selection procedure | selectCox |  | selectFGR |  | crrp |  |
| --- | --- | --- | --- | --- | --- | --- |
| Covariable (baseline value) | relapse | death | relapse | death | relapse | death |
| Donor age (10y increase) | - | 1.1 (1.10;1.12)<br>0.009 | - | - | - | - |
| Patient age (10y increase) | - | 1.4 (1.41;1.43)<br><0.001 | 0.92 (0.91;<br>0.92)<br><0.001 | 1.5 (1.4; 1.5)<br><0.001 | 0.96 (0.95; 0.96)<br><0.001 | 1.3 (1.29; 1.31)<br><0.001 |
| Donor: Others | - | 0.7 (0.5;0.9)<br>0.003 | - | - | - | - |
| Others than Male patient,<br>female donor | - | - | - | - | - | - |
| CMV match | - | - | - | - | - | - |
| First transplant | - | 1.9 (1.0;3.7)<br>0.056 | - | - | - | - |
| Karnofsky Performance<br>Status: good | - | 1.9 (1.5, 2.3)<br><0.001 | - | - | - | 1.5 (1.3;1.7)<br><0.001 |
| Disease stage (0: early, 1:<br>intermediate, 2: advanced) | 1.4 (1.3;1.5)<br><0.001 | 1.3 (1.2 ;1.4)<br><0.001 | 1.3 (1.2; 1.4)<br><0.001 | 1.3 (1.2;1.4)<br><0.001 | 1.2 (1.2;1.3)<br><0.001 | 1.1 (1.1;1.2)<br><0.001 |
| Non Myeloablative<br>conditioning | - | - | - | - | - | - |
| TBI: no | - | - | - | - | - | - |
| Antithymocyte globulin<br>(ATG): no | - | 0.8 (0.7; 0.9)<br>0.009 | - | - | - | - |

Discrimination is measured with the cumulative/dynamic AUC until the median follow up time: 0.75 years, estimated with Rubin’s rule over 10-fold cross validation and for each endpoint. For *selectCox*, AUC were 58% (95%CI 50-66) and 66% (95%CI 52-80) for relapse and death respectively. For *selectFGR*, AUC were 59% (95%CI 49-70) and 65% (51-80) for relapse and death respectively. For *crrp*, AUC were 59% (95%CI 49-70) and 67% (52-82) for relapse and death respectively.

The estimated AUC for all models is above 50%. This means, that the model assigns the higher predicted risk to a patient who experience the event of interest within 0.75 years with the reported probability compared to a patient experiencing the other event or is censored until 0.75 years. This discrimination is better for endpoint death, but all three variable selection procedures and the two model specifications (CSH, SDH) seems to discriminate comparably.

## 5. Discussion

A number of lessons can be learned from these simulations. First, when using available methods for performing variable selection in the presence of competing risks, the true data generating model is rarely identified. This is of high importance since the main purpose of building a multivariable model in epidemiological studies is often to identify risk factors adjusted for some confounders. As is the case for other regression models (e.g., linear or logistic), the resulting model after variable selection is very likely to miss some associated variables and/or to include wrongly associated variables. Interestingly, predictions based on the selected model are only slightly less accurate than those from the true model. Even if the true model is almost certainly not identified, it seems that missing variables (or additional variables) have a marginal impact on prediction. Therefore, if the selection procedure aims at building a predictive tool, all proposed methods could be used. This fact has already been noticed in the literature in other regression context (e.g., logistic in (25)).

A consensus has emerged for the analysis and reporting of competing risk: both CSH and SDH ratios should be reported to fully understand the effect of variables on the competing outcomes (9). However, some variable selection methods only focus on the event of interest for SDH, which may seem inconsistent in understanding the effect of selected variables on the other SDH, and both CSH.

It is worth noting that, for each procedure, using options different from the default options proposed by the package author may dramatically change the performance of the selection strategy. In this work, we highlighted the fact that the fast backward algorithm yields a higher false discovery rate than the naïve iterative approach. This means that the end user of a package should also care about the implementation choices of the package author, which can be difficult in practice for non-specialists. Namely, one should not make the conclusion that selection on the subdistribution hazard of the event of interest only is associated with a high FDR since our data does not support this claim.

This sensitivity of the model with respect to the analysis method and its implementation is also striking in the real data application, for which the proposed models are very different depending on the method used. However, the discrimination and calibration performances seem to be rather stable, consistently with the simulation study.

Our study has some limitations. First, in our simulation, covariates were independently drawn from a normal or binomial distribution. In real data however, correlation between predictors is frequently observed and we would expect selection algorithms to perform even worse in this situation. We also limited our covariates to be either binary or quantitative. In real life however, categorical variables with more than two levels are common, and the distribution across modalities may also impact the performances of selection algorithms, e.g. if some modalities have low sample size. Moreover, the choice of AUC for evaluating model predictions can be challenged. In some cases where probabilities are close, predictions could be near perfect while the AUC remain modest. This situation can occur depending on values of baseline hazards and effect size of covariates (26). On the contrary, in some applications, only a good calibration is required. However, AUC is still often used as a discrimination measure for model evaluation in the literature. Another limitation comes from the scope of the paper which is limited to regression models, thus excluding Machine Learning (ML) methods as Random survival forests or boosting. However, a recent review (27) is available and deals with the application of ML methods for competing risks. Finally, as is the case for all simulation studies, the results might not be generalizable to particular settings (e.g. high dimensionality, where cross-validation is often used to avoid overfitting issues), although the proposed simulation scenario is rather complex and comprehensive, as compared to other published papers.

Based on these results, we would like to make some recommendations for analysts. First, (1) acknowledge as a limitation the fact that the identified multivariable model probably includes variables not associated with the outcome of interest, or miss truly associated variables, (2) report the number of events per variable corresponding to the risk under consideration, and (3) consider using multiple selection strategies to mitigate the risk of an implementation issue.

## Data Availability

All data produced in the present study are available upon reasonable request to the authors

## 6. Declarations

### Ethics approval and consent to participate

Data presented in section 4 (application) originated from the German registry for stem cell transplantation (24), which has been conducted in accordance with the Declaration of Helsinki, and which received approval 226/17 and 141/19 from the ethical board of the Ulm University (Clinical trial number: not applicable). Informed consent had been retrieved prior to reporting.

### Consent for publication

Not applicable

### Availability of data and materials

The datasets generated and analyzed during the current study are available at https://github.com/jmullaert/VariableSelection

### Authors contributions

JM was a major contributor in conducting the simulations writing the manuscript. SS performed and reported the analysis of section 4. PA and AL directed the work and substantively revised the manuscript. All authors read and approved the final manuscript.

## Acknowledgement

The authors gracefully thank Jan Beyersmann for discussing the manuscript and the German registry for stem cell transplantation (DRST) for providing the data used in this paper.

## Supplementary materials

**Supplementary table S1:** Description of the parameters used for the simulation under proportional cause-specific hazard. Censoring rates refers to the parameter of the exponential distribution used to generate censoring times.

| Scenario | Baseline hazard<br>for risk 1 ($h_{10}$) | Baseline hazard<br>for risk 2 ($h_{20}$) | Censoring rate |
| --- | --- | --- | --- |
| **S1** | 0.04 | 0.04 | 0.01 |
| **S2** | 0.04 | 0.01 | 0.01 |
| **S3** | 0.04 | 0.04 | 0.001 |
| **S4** | 0.04 | 0.01 | 0.001 |

**Supplementary table S2:** Description of the parameters used for the simulation under proportional subdistribution hazard. Censoring rates refers to the parameter of the exponential distribution used to generate censoring times.

| | Asymptotic cumulative<br>incidence of event 1 for<br>$X = 0$ | Rate of<br>occurrence<br>of event 2 | Censoring rate |
| --- | --- | --- | --- |
| Scenario | ($p$) | ($\lambda_0$) | |
| **S1** | 0.8 | 0.1 | 0.7 |
| **S2** | 0.5 | 0.1 | 0.7 |
| **S3** | 0.8 | 0.1 | 0.2 |
| **S4** | 0.5 | 0.1 | 0.2 |

**Supplementary figure S3:**
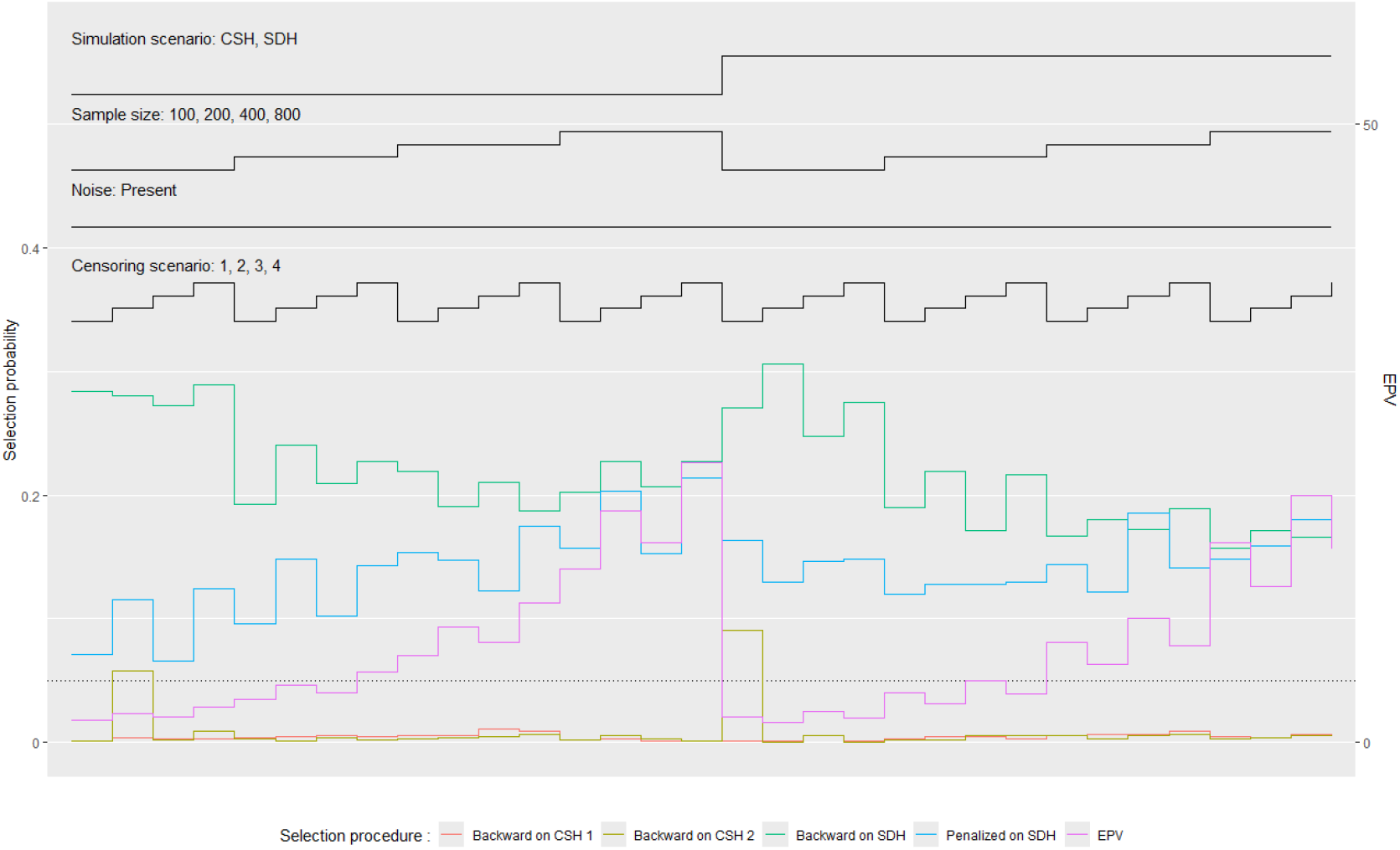
False discovery rate (left axis) and EPV (right axis) for selection procedures under the alternative hypothesis according to sample size, censoring scenario and simulation scenario (32 scenarios). Only noise variables are considered here to avoid issue with model misspecification.

